# Phenotypic structure and genetic correlates of adolescent psychiatric symptomatology in versus out of autism

**DOI:** 10.64898/2026.10.15.26365562

**Authors:** Taylor R. Thomas, Susan S. Kuo, C. Hope Lefebvre, Jing Zhang, Robert Havkin, F. Kyle Satterstrom, Jack M. Fu, Chiara Auwerx, Celia van der Merwe, Karmel W. Choi, Michael E. Talkowski, Gagan Joshi, Elise B. Robinson

## Abstract

**Objective:** Mental health challenges are highly prevalent in autism, but the degree to which they are similar in experience and etiology to the general population remains unclear. This study’s objective was characterization of phenotypic and genetic architectures of psychiatric symptomatology in versus out-of-autism.

**Methods:** Psychiatric symptoms were evaluated using the Child Behavior Checklist (CBCL) in 22,152 children (8-15 years): 11,111 autistic children with verbal abilities from SPARK and 11,041 non-autistic children from ABCD. The CBCL’s canonical, population-wide factor structure and genetic associations were compared between cohorts. In parallel, exploratory factor analysis was conducted in SPARK, with resulting dimensions associated with key clinical phenotypes (cognitive ability, co-occurring neuropsychiatric diagnoses), polygenic scores (PGS) for psychiatric disorders, and de novo burden in autism-associated genes.

**Results:** Confirmatory factor analysis supported the canonical structure in ABCD but not in SPARK. Psychiatric PGS associations with canonical CBCL subscales were weaker in SPARK than in ABCD, with the ADHD PGS association to Attention problems lower in SPARK (β=0.05 [CI: 0.04,0.06] than in ABCD (β=0.17 [0.13,0.21]). SPARK exploratory factor analysis replicated canonical internalizing factors while revealing other structural departures, including a distinct Developmental Delays factor and dispersion of social problems across multiple factors. The Developmental Delays factor had the strongest associations with de novo burden and lower cognitive ability but was weakly associated with psychiatric PGS. In contrast, common genetic variation explained larger proportions of variances in somatic and inattentive factors and had stronger PGS associations.

**Conclusions:** Correlation structures and genetic architectures of psychiatric symptoms differed in autism compared to the broader population, emphasizing the need for autism-specific approaches in assessment and treatment.

## Introduction

Autistic individuals face a disproportionate burden of mental health problems. Compared to people without autism diagnoses, autistic individuals show increased risk for many neuropsychiatric disorders, including ADHD, obsessive-compulsive disorder, substance use disorder, anxiety, depression, bipolar disorder, and schizophrenia [1–3]. Suicide risk is also markedly elevated in the autism community, particularly among autistic individuals without intellectual disability [4].

Characterization of mental health challenges among autistic individuals is complicated by autism’s highly heterogeneous clinical presentations [5,6]. Among the foundational challenges in autistic mental health care is a lack of psychiatric symptom assessments specifically validated in autistic populations. Absent valid symptom-level assessments, individual differences and changes in mental health can only be tracked at the diagnostic level, obscuring estimates of prognosis and treatment response. A better understanding of psychiatric symptom measurement in autism could have immediate clinical implications and form a basis for future research.

One of the most widely used assessments of psychiatric symptoms in children is the Child Behavior Checklist [7] (CBCL). The CBCL is a 119-item questionnaire that captures a breadth of psychiatric symptoms, historically organized into eight canonical subscales: Aggressive and Rule-Breaking (Externalizing); Anxious, Withdrawn, and Somatic (Internalizing); and Attention, Social, and Thought problems. Several confirmatory factor analyses in smaller samples have suggested that this general population-normed canonical structure fits poorly in autism [8–10]. This calls into question the comparability of psychiatric symptom dimensions between autistic and non-autistic samples. It also complicates the etiologic study of mental health symptomatology in autistic populations. The phenotypic and genetic architectures of psychiatric symptomology in autism have not yet been investigated at scale, nor have they been compared to those same architectures in a non-autistic population.

To identify a stable factor structure for psychiatric symptoms in autism, and to etiologically characterize these symptoms across autistic and non-autistic populations, we leveraged the Simons Foundation Powering Autism Research study [11] (SPARK; N=11,111) as an autism cohort and the Adolescent Brain Cognitive Development study [12] (ABCD; N=11,041) as a whole population, non-autistic cohort. We found the CBCL’s canonical eight-factor structure to be a weaker representation of psychiatric symptom dimensions in SPARK compared to ABCD. We instead derived a seven-factor CBCL structure in SPARK that indexed alternative dimensions of psychiatric symptoms and developmental delays. The developmental delays factor had the strongest associations with de novo variant burden and lower cognitive ability but was weakly associated with psychiatric PGS. In contrast, common genetic variants explained larger proportions of phenotypic variances for inattentive and somatic factors and had stronger associations with psychiatric PGS. Altogether, our study demonstrates feasibility of large-scale psychometric validation in autism, utility of a refactored psychiatric symptom assessment in autism, and specificity of the relationships between certain psychiatric symptoms in autism.

## Methods

### Study Participants and Phenotypes

Ethical approval for this study was obtained from the Massachusetts General Brigham Institutional Review Board under the study name Molecular Study of Cognitive and Behavioral Variation (IRB: 2015P002376). Study participants included a total of 22,152 children from SPARK and ABCD (Figure 1, Table S1). Both cohorts were recruited from several locations across the United States. We restricted our SPARK sample to match the same age range as ABCD, which was 8-15 years. We further restricted SPARK to participants with verbal abilities due to known CBCL biases in non/minimally verbal individuals [13,14]. We defined verbal ability as both sentence-level speech at the most recent parent-reporting and positive endorsement of the first item in the Social Communication Questionnaire that assesses use of short phrases or sentences [15]. We excluded ABCD participants with diagnoses of autism, schizophrenia, cognitive impairment, or epilepsy. Both studies recruited participants from multiple locations in the United States, with the Area Deprivation Index [16] (ADI) national percentile ranging from 1-100 in both cohorts. The ADI mean difference (MD) was 7.2 [6.4,7.9]; p<0.001, with the SPARK average of 47.0 (SD=25.9) and the ABCD average of 39.9 (SD=26.9), suggesting reasonable socioeconomic representation across cohorts. Cognitive impairment status in SPARK was defined from the parent’s most recent report on whether the participant had been diagnosed with cognitive impairment or intellectual disability. Co-occurring neuropsychiatric diagnoses in SPARK were parent-reported professional diagnoses from the background medical history questionnaire, in which we included participants who were at least six years old at reporting (N=9,517). Psychiatric diagnoses in ABCD were assessed using the Kiddie Schedule for Affective Disorders and Schizophrenia [17] (KSADS) at baseline intake (N=11,041).

**Figure 1.**
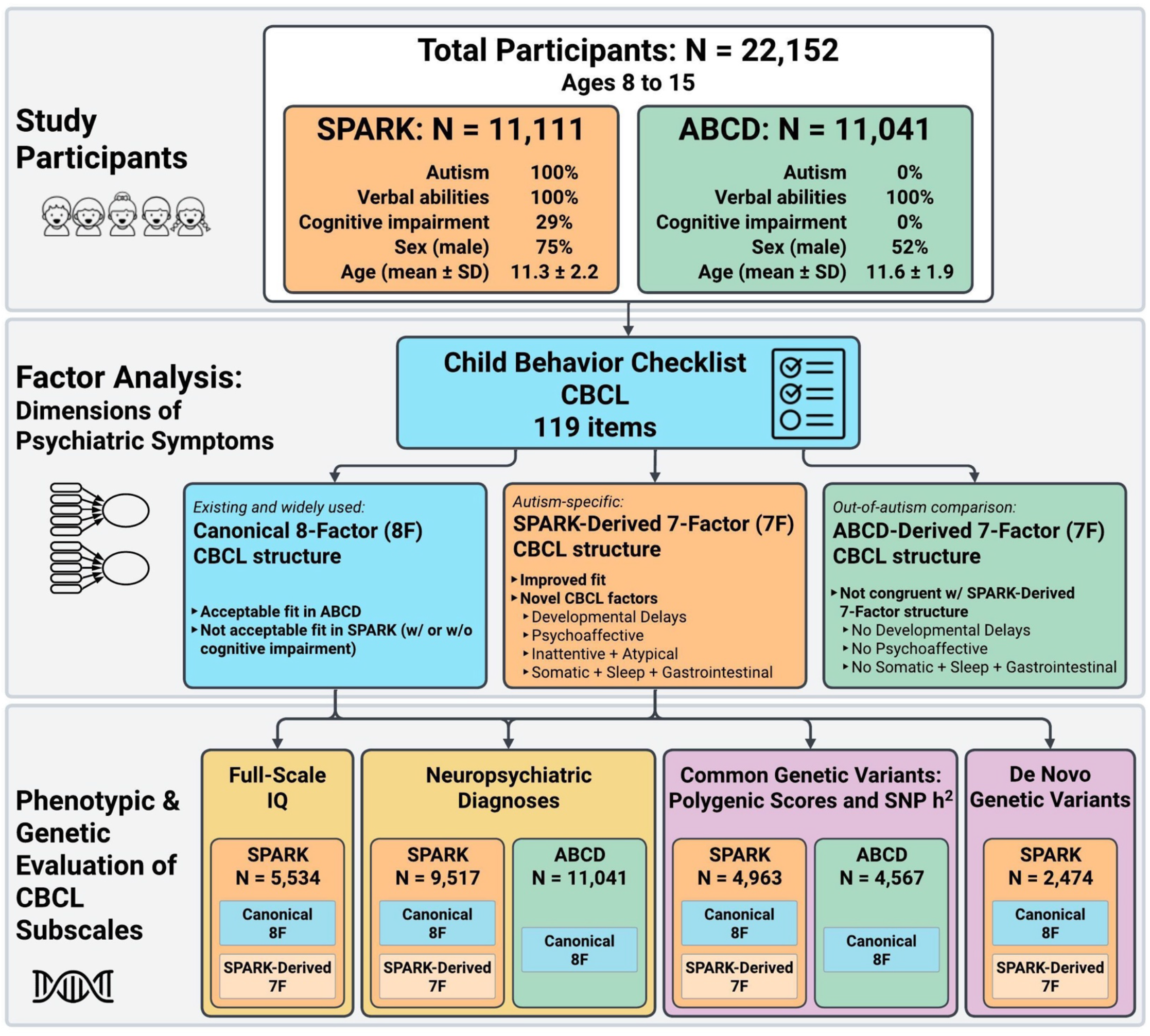
Overview of Study Participants and Analyses. See Table S1 for additional demographics.

### Factor Analyses

As all CBCL items are on a 3-point Likert scale, we used factor analysis methods with ordinal specifications. Confirmatory factor analyses (CFAs) were conducted using lavaan [18] with the weighted least squares mean-and-variance-adjusted estimator. CBCL item means and variances were higher in SPARK (average mean=0.52; average variance=0.40) than in ABCD (average mean=0.14; average variance=0.14; Table S4). We removed the following five items that had near-zero variance in both cohorts: drinks alcohol, uses tobacco, uses drugs, sets fires, and sexual problems. With the remaining 114 items, we used three methods to determine the optimal number of factors to extract in SPARK: 1) variance explained from principal component analysis of the correlation matrix, 2) scree regression [19], and 3) sequential exploratory factor analysis (EFA) from 1-15 factors (Table S5). We determined a seven-factor solution to be optimal based on each component explaining >2% of the variance, 47% cumulative variance explained and a scree R^²^ of 0.77 (recommended cutoff ≥0.75). EFAs were conducted using psych [20] with Promax rotation, weighted least squares factoring method, and polychoric correlations. We compared EFA structures using the Tucker congruency coefficient method [21], in which φ≥0.95 indicates nearly identical factors.

The final SPARK-derived EFA was converted to CFA structure and item-summed subscales using a loading threshold of ≥0.3 (Figure S2). Due to the possibility of an item having more than one meaning and/or a different meaning in the context of autism (e.g., running away from home potentially corresponding to either rule-breaking or elopement in autism due to sensory overwhelm [22]), we allowed items to cross-load to more than one factor. The final SPARK-derived factor structure leveraged 103 items, 20 that cross-loaded to two factors and one that cross-loaded to three factors (Table S7).

### Genetic Data

Many details of our genetic data quality control, imputation, merging, and polygenic score (PGS) workflows have been previously published [23] and can be found in full in the Supplement. We tested PGS associations with the CBCL subscales separately in each cohort (SPARK N=4,963; ABCD N=4,567; Table S11) and tested for PGS-by-cohort interaction effects. PGS were calculated using LDpred2 [24,25] from genome-wide association studies of cognitive performance [26] and case-control studies of four major psychiatric disorders: autism [27], ADHD [28], depression [29], and schizophrenia [30]. We analyzed de novo burden associations in the subset of SPARK with complete trios (N=2,474; Table S12). We defined de novo burden as carriers of a de novo protein-truncating variant or de novo coding copy-number variant in an autism-associated gene (253 genes), or carriers of a de novo genomic disorder deletion or duplication associated with autism or intellectual disability (e.g., 22q11.2 deletion). The de novo calls and autism-associated gene list are from the latest unpublished exome sequencing aggregation effort by the Autism Sequencing Consortium [31].

### Statistical Analyses

All phenotypic and genetic associations with the CBCL subscales included sex and age at CBCL assessment as covariates. Regression coefficients are standardized estimates with 95% confidence intervals (CI) in brackets. We used logistic regression for case/control diagnosis outcomes, ordinal logistic regression for full-scale IQ outcomes, and negative binomial regression for CBCL subscale outcomes. For t-tests and correlations, CBCL subscales were adjusted for age and sex with negative binomial regression residualization and then standardized. P-values were adjusted for multiple comparisons using the false discovery rate method unless stated otherwise. P-values were adjusted separately per cohort and CBCL subscale group (i.e., canonical and SPARK-derived) because of redundancy between subscale groups and to maintain the same adjustment level across cohorts.

## Results

### Assessing the Canonical, Population-Normed CBCL Subscales in SPARK and ABCD

SPARK had higher means and variabilities for all eight canonical CBCL subscales compared to ABCD; these patterns were consistent across SPARK with and without cognitive impairment (Figure 2A, Table S2). The widely used T-scores employ critical cutoffs (two SD above the normed mean) that indicate clinically significant psychiatric symptoms suggestive of a specific psychiatric condition. SPARK had higher rates of critical T-scores across all subscales (Table S1). Differences between SPARK and ABCD were most prominent for Thought problems, with 58% of SPARK at or above the critical threshold compared to 3% of ABCD.

**Figure 2.**
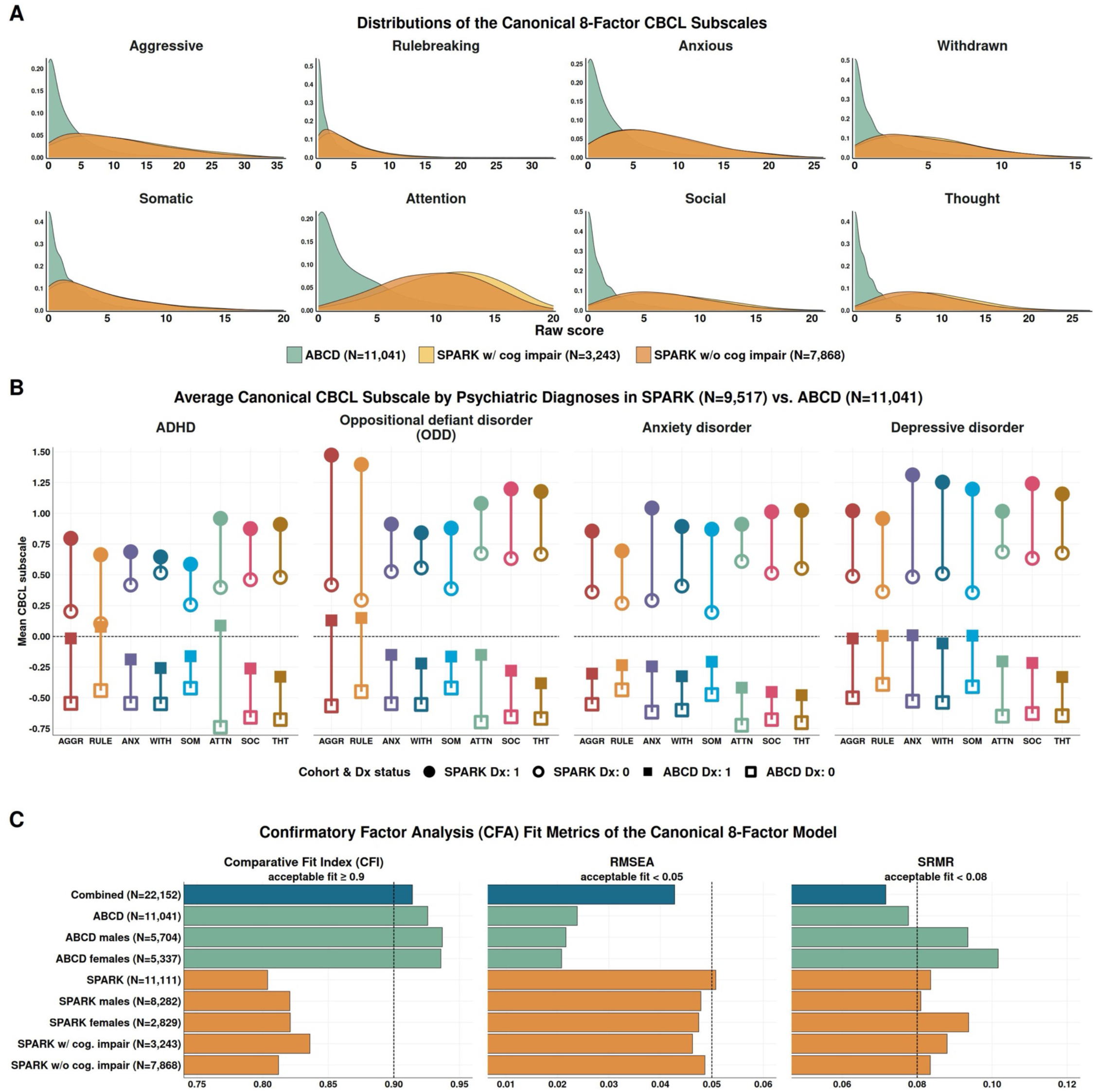
Comparison of the Canonical Eight-Factor CBCL Subscales in SPARK and ABCD. **A**. Distribution of the canonical CBCL subscales stratified by autism/cohort (SPARK versus ABCD)and cognitive impairment within SPARK. **B**. Average CBCL subscales by cohort and co-occurring ADHD, ODD, anxiety, and depressiondiagnoses. The subscales were adjusted for age and sex with negative binomial regressionresidualization and then standardized. **C**. CFA fit metrics for the canonical eight-factor structure in SPARK and ABCD subgroups.

The ADHD prevalence was higher in SPARK (58%) than in ABCD (14%), whereas prevalences were comparable for oppositional defiant disorder (ODD; SPARK=12%; ABCD=14%), anxiety (SPARK=38%; ABCD=33%), and depression (SPARK=11%; ABCD=6%). ADHD was associated with higher Attention scores in SPARK (MD=0.56 [0.53,0.59]) and ABCD (MD=0.83 [0.79,0.87]), ODD with higher Aggressive scores in SPARK (MD=1.06 [0.99,1.13]) and ABCD (MD=0.70 [0.65,0.74]), and anxiety with higher Anxious scores in SPARK (MD=0.75 [0.71,0.79]) and ABCD (MD=0.37 [0.35,0.40]) (Figure 2B; Table S3).

Despite not having a given neuropsychiatric diagnosis, the vast majority of SPARK subsets *without* the co-occurring diagnosis had higher CBCL scores than ABCD subsets *with* the diagnosis. As an example, SPARK without ADHD had significantly higher Attention scores than ABCD with ADHD (MD=0.31 [0.27,0.36]; t=13.30; p<0.001). Furthermore, the effect of a one-point increase in a CBCL subscale was lower in SPARK than in ABCD: the Attention subscale effect for an ADHD diagnosis in SPARK was OR=1.20 [1.19,1.22], whereas in ABCD the effect was OR=1.43 [1.40,1.45]. At least two non-mutually exclusive explanations beyond diagnostic imprecision may be contributing to these patterns. First, some symptomatic presentations of autism phenocopy other psychiatric conditions, producing significantly elevated psychiatric symptoms without true presence of the co-occurring condition. Second, diagnostic overshadowing raises the clinical threshold for recognizing and diagnosing elevated psychiatric symptoms as a psychiatric condition in autistic individuals. Separating these mechanisms requires etiological studies, particularly genetic approaches.

To interrogate how well the canonical eight-factor structure represents underlying CBCL correlations in versus out of autism, we conducted CFAs stratified by autism, sex, and cognitive impairment (Figure 2C; Table S6) and tested for measurement invariance between cohorts. While the canonical structure had an acceptable fit in ABCD (comparative fit index (CFI)=0.926; root mean square error of approximation (RMSEA)=0.024; standardized root mean squared residual (SRMR)=0.078), it had a weaker fit in SPARK (CFI=0.804; RMSEA=0.051; SRMR=0.084). Measurement invariance tests further indicated non-equivalent factor structures between cohorts, with a baseline CFI of 0.914 that dropped to 0.854 upon testing whether the form of the factor structures were equivalent between cohorts.

### Identifying an Alternative CBCL Factor Structure in SPARK

To identify a better-fitting structure in the context of autism, we conducted EFA of the CBCL itemset in SPARK. Unlike CFA, which is used to assess an established factor structure, EFA is used to determine a factor structure in a given itemset without known priors. We used three methods to determine the number of factors and identified seven factors to be optimal (Table S5). This seven-factor model fit better than a general psychopathology factor model with seven factors (ω^h^=0.77). To confirm stability of the seven-factor structure in SPARK, we randomly split the SPARK sample and compared factor structures across groups. The split-half structures mirrored each other, with an overall congruency of φ=0.97 and per-factor congruencies ranging from 0.91-0.99 (Figure S1), indicating all factors were nearly identical. To compare the SPARK-derived seven-factor structure to an out-of-autism seven-factor structure, we extracted seven factors in ABCD. The SPARK-derived versus ABCD-derived structures showed moderate congruency, with an overall congruency of 0.87 and per-factor congruencies ranging from 0.19-0.97 (Figure 3A).

**Figure 3.**
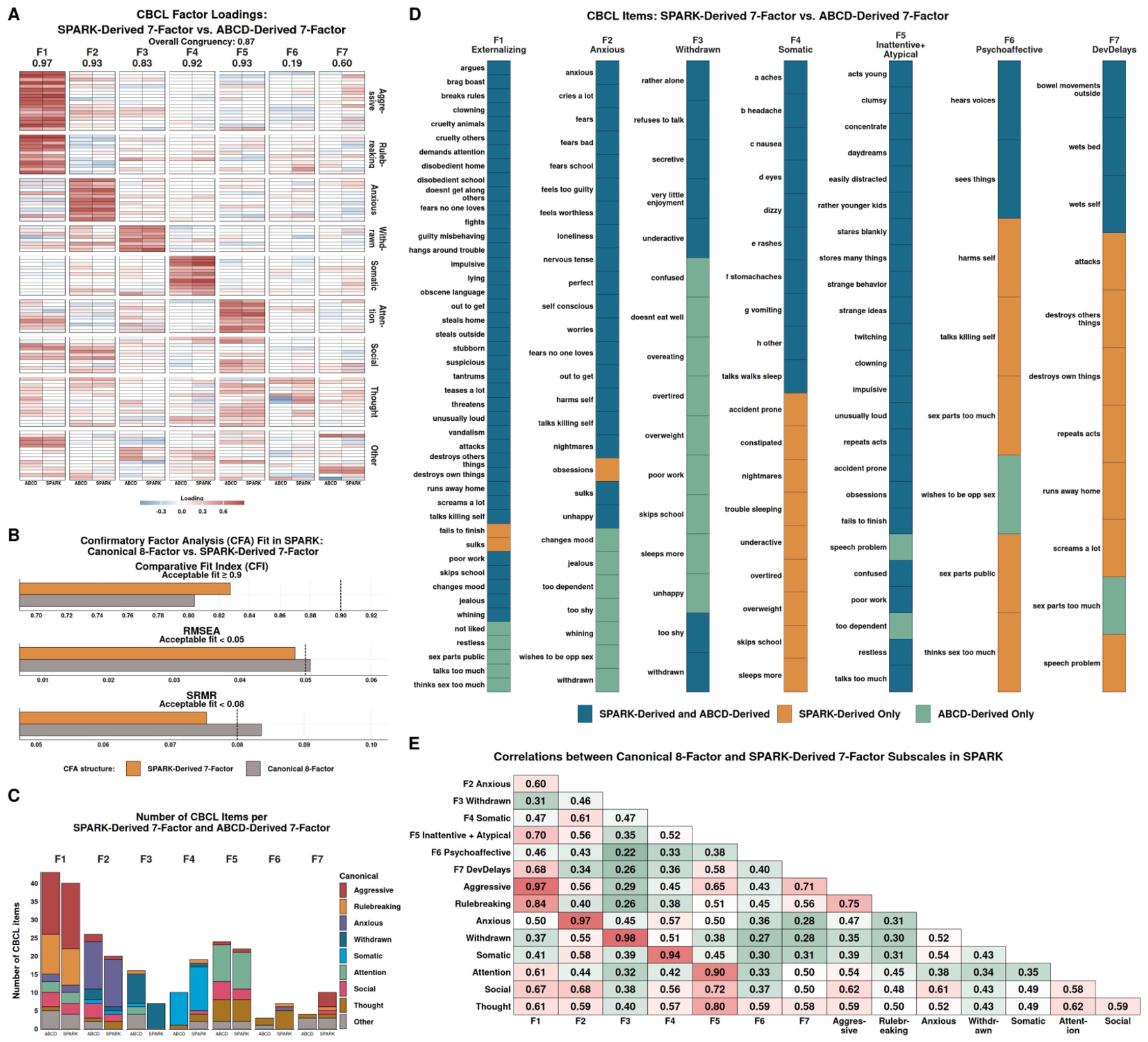
Identifying an Alternative CBCL Factor Structure in SPARK. **A**. Factor loadings from the seven-factor EFAs in SPARK and ABCD and the congruencies (φ) between factors. The breaks along the y-axis indicate the canonical CBCL structure. EFA loadings ≥0.3 were included as latent variables in the final seven-factor structures for CFA. **B**. Canonical eight-factor and SPARK-derived seven-factor CFA fit metrics in SPARK. **C**. CBCL item counts by SPARK-derived and ABCD-derived seven-factor structures, with the item color indicating that item’s canonical eight-factor assignment. **D**. List of CBCL items for each SPARK-derived and ABCD-derived factor, with the item color indicating whether that item is unique to the SPARK-derived or ABCD-derived structure. **E**. Correlations between the canonical and the SPARK-derived subscales in SPARK. The subscales were adjusted for age and sex prior to calculating correlations.

Reflecting extensive heterogeneity of psychiatric symptoms in autism, the SPARK-derived seven-factor structure yielded a slightly improved CFA fit in SPARK compared to the canonical structure (CFI=0.827; RMSEA=0.048; SRMR=0.075; Figure 3B). Three SPARK-derived factors broadly recaptured some of the canonical factors (Figure 3C-3E, Table S7-S8). Externalizing Factor-1 was highly correlated with canonical Aggressive (r=0.97) and Rulebreaking (r=0.84), Anxious Factor-2 with canonical Anxious (r=0.97), and Withdrawn Factor-3 with canonical Withdrawn (r=0.98).

However, items from other subscales, most notably canonical Social items, were dispersed across several SPARK-derived factors. Somatic-Dysregulation Factor-4 captured somatic complaints, sleep and energy problems, and gastrointestinal complaints. Inattentive-Atypical Factor-5 captured problems of obsessiveness, failure to concentrate, and strange behaviors and ideas. SPARK-derived Psychoaffective Factor-6 was distinct from ABCD-derived Factor-6 (φ=0.19) and captured problems of inappropriate sexual behaviors, suicidal ideation, self-harm, and auditory and visual hallucinations, suggesting a psychosis and mood factor. SPARK-derived Developmental Delays Factor-7 was also distinct from the canonical structure and ABCD-derived Factor-7 (φ=0.60) and captured problems with incontinence, repeating acts, running away from home, screaming, and speech problems.

### Phenotypic Profiles of the CBCL Subscales in SPARK

We assessed relationships between CBCL subscales and full-scale IQ in SPARK (N=5,534; Figure 4A; Table S9). Lower full-scale IQ was most strongly associated with Developmental Delays Factor-7 (β=−0.56 [−0.61,−0.51]). In contrast, higher full-scale IQ was associated with Anxious Factor-2 (β=0.11 [0.06,0.15]). We also assessed relationships between SPARK-derived subscales and co-occurring psychiatric disorders (SPARK N=9,517; Figure 4B; Table S10). In addition to the four psychiatric disorders, we included four developmental and neurological disorders: cognitive impairment (prevalence of 30%), language delay or disorder (40%), motor delay or developmental coordination disorder (16%), and seizures/epilepsy (5%). Developmental Delays Factor-7 was the strongest predictor of cognitive impairment (β=0.47 [0.42,0.51]), language delay/disorder (β=0.33 [0.29,0.37]), motor delay/disorder (β=0.40 [0.35,0.46]), and epilepsy/seizures (β=0.43 [0.35,0.51]). Developmental Delays Factor-7 was the weakest predictor of anxiety (β=0.21 [0.17,0.25]) and depression (β=0.17 [0.11,0.24]), whereas Anxious Factor-2 was the strongest predictor of anxiety (β=0.74 [0.69,0.79]) and depression (β=0.79 [0.72,0.85]).

**Figure 4.**
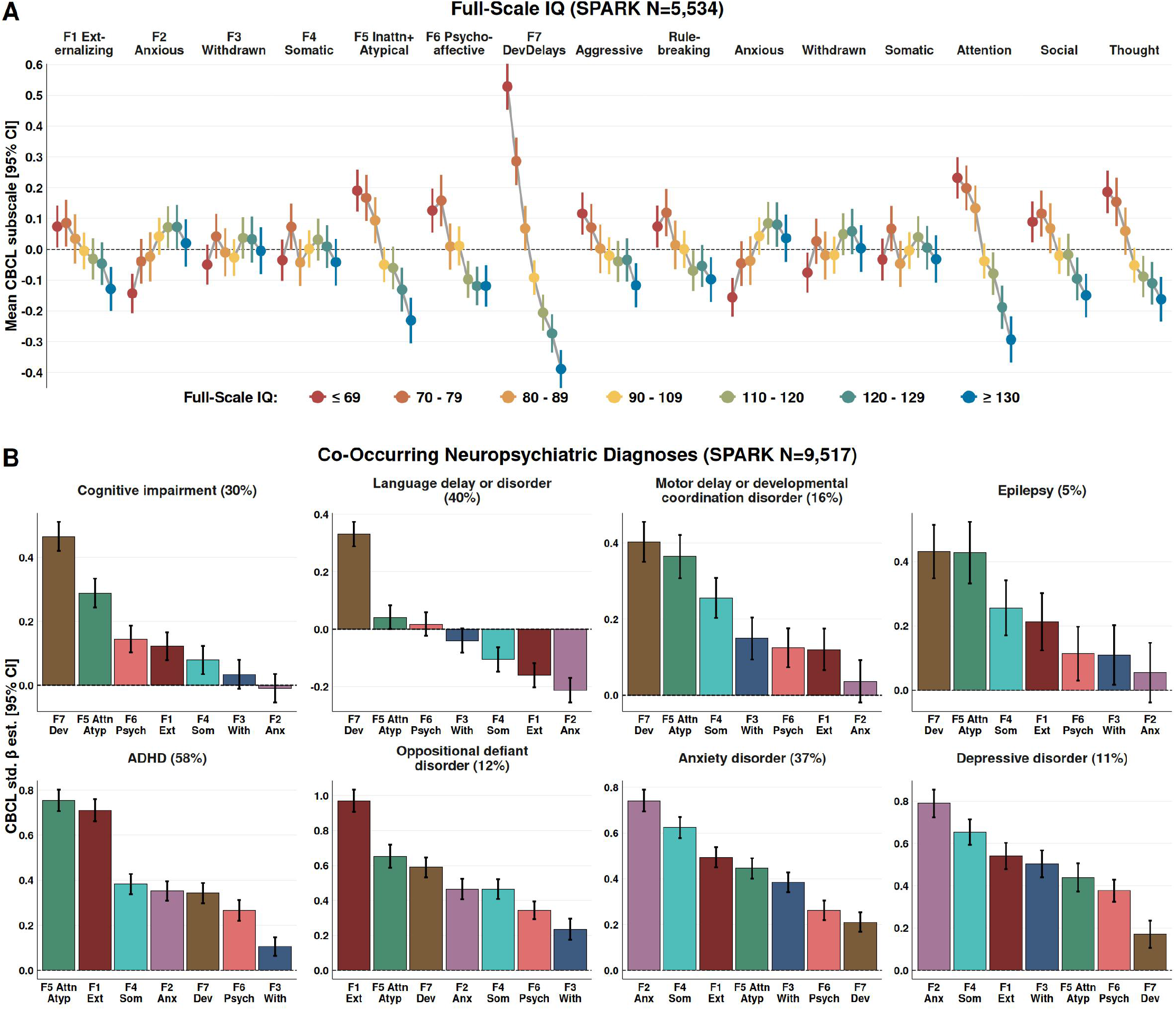
Phenotypic Profiles of the CBCL Subscales in SPARK. **A**. CBCL associations with full-scale IQ in SPARK. **B**. CBCL associations with eight co-occurring neuropsychiatric diagnoses in SPARK. The CBCL β estimates with 95% CI are from logistic regression predicting each co-occurring diagnosis:Logistic regression: *Diagnosis[1] ~ CBCL Subscale + Age + Sex*

### Genetic Associations with the CBCL Subscales in SPARK and ABCD

We estimated the proportion of each subscale’s phenotypic variance explained by common genetic variation (i.e., SNP-based heritability or h²; Table S13) and compared CBCL subscale associations with psychiatric PGS (Figure 5A; Table S14). Common genetic associations, in general, were stronger in ABCD (N=4,567) than in SPARK (N=4,963). The strongest PGS-by-cohort interaction was the ADHD PGS with the Attention subscale (Z=−7.88, p<0.001), in which the ADHD PGS effect was higher in ABCD (β=0.17 [0.13,0.21]) than in SPARK (β=0.05 [0.04,0.06]), as was the Attention h² (ABCD h²=0.22; SE=0.08; p^nominal^=0.002, SPARK h²=0.09; SE=0.07; p^nominal^=0.117). The depression PGS interaction on the Social subscale was significant (Z=−6.39; p<0.001), again in which the PGS effect was higher in ABCD (β=0.18 [0.13,0.22]) than in SPARK (β=0.05 [0.04,0.07]), and the h² was higher in ABCD (h²=0.13; SE=0.08; p^nominal^=0.036) than in SPARK (h²=0.06; SE=0.07; p^nominal^=0.201). The autism PGS was not associated with any subscales in SPARK but was associated with six subscales in ABCD, with the strongest association with the Attention subscale (β=0.08 [0.04,0.12]).

**Figure 5.**
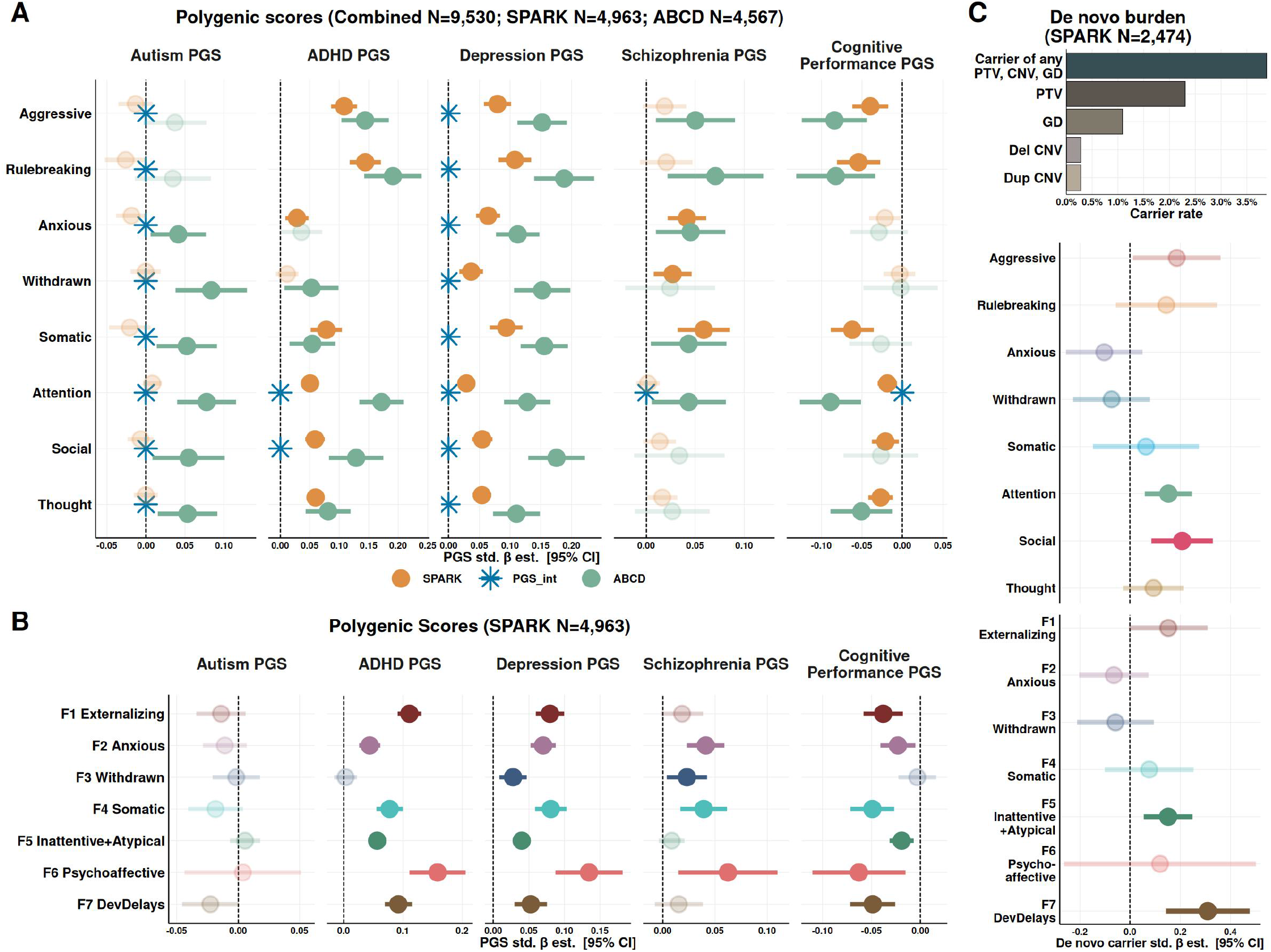
Polygenic and De Novo Genetic Associations with the CBCL Subscales. **A**. PGS associations with the canonical eight-factor subscales. PGS were tested separately percohort and in a combined-cohort model with a PGS-by-cohort interaction term:Negative binomial regression: *CBCL Subscale ~ PGS + Cohort + (PGS × Cohort[SPARK]) + Age+ Sex*ignificant PGS-by-cohort interaction effects are indicated with blue asterisks. **B**. PGS associations with the SPARK-derived subscales in SPARK.Negative binomial regression: *CBCL Subscale ~ PGS + Age + Sex* **C**. De novo burden associations with the canonical and SPARK-derived subscales in SPARK.Negative binomial regression: *CBCL Subscale ~ De Novo Burden Carrier + Age + Sex*

We analyzed PGS associations with the SPARK-derived subscales in SPARK (Figure 5B). Here, the depression PGS was most strongly associated with Psychoaffective Factor-6 (β=0.13 [0.09,0.18]), as was the schizophrenia PGS (β=0.06 [0.01,0.11]). The subscale with the highest h² was Externalizing Factor-1 (h²=0.29; SE=0.07; p^nominal^<0.001), which was most strongly associated with the ADHD PGS (β=0.11 [0.09,0.13]). For Inattentive-Atypical Factor-5, the ADHD PGS had the strongest association (β=0.06 [0.04,0.07]) and the h² was 0.15 (SE=0.07; p^nominal^=0.017).

Lastly, we examined associations with de novo burden in the SPARK subset with complete trios (N=2,474; Figure 5C). The overall de novo burden carrier rate was 3.9%, in which 2.3% carried a protein-truncating variant, 0.6% carried an exonic copy-number variant, and 1.1% carried a genomic disorder variant. De novo variant burden was most strongly associated with SPARK-derived Developmental Delays Factor-7 (β=0.31 [0.14,0.48]) and to a lesser extent Inattentive-Atypical Factor-5 (β=0.15 [0.05,0.25]). For the canonical subscales, de novo burden was associated with Attention (β=0.15 [0.06,0.25]) and Social (β=0.21 [0.08,0.33]).

## Discussion

Autistic individuals experience many psychiatric symptoms at higher rates than the general population, yet their symptom profiles are often underrecognized as diagnosable mental health conditions. Across four neuropsychiatric disorders that frequently emerge by childhood (i.e., ADHD, ODD, anxiety, and depression), rates of associated psychiatric symptoms were significantly elevated among autistic children compared to non-autistic children. Autistic children without a given neuropsychiatric diagnosis showed higher rates of associated psychiatric symptoms compared to non-autistic children with the diagnosis. This suggests that a substantial proportion of autistic children struggle with mental health challenges at levels that would warrant diagnosis and further care in non-autistic children. Addressing this possible healthcare inequity begins with improving the measurement and tracking of psychiatric symptoms in autism.

Patterns of co-occurring psychiatric symptoms in SPARK differed from patterns in ABCD. We observed a lack of measurement invariance for the CBCL’s canonical structure between SPARK and ABCD, and we identified an alternative structure that better explains psychiatric symptom patterns in SPARK. CBCL structural differences between cohorts emphasize the importance of integrating developmental and behavioral patterns to gauge psychiatric symptomology in autism. The findings may also indicate differential clinical meaning of certain questionnaire items, suggesting that similar observed behaviors do not necessarily represent equivalent underlying clinical constructs across autistic and non-autistic children. Some behaviors captured by CBCL items, including psychosis-like experiences [32,33], may reflect psychiatric morbidity but also sensory overload [34,35], rigidity, communication barriers, or developmental level. Among the seven SPARK-derived factors, developmental delays were concentrated in one dimension, while social symptoms were dispersed across multiple dimensions. Elevated burdens in other psychiatric dimensions did not track with higher burdens in developmental delays. This suggests that developmental delays may be relatively independent from other psychiatric symptoms in autism and/or that psychiatric symptoms are not adequately understood and measured in autistic individuals with higher burdens of developmental delays [36,37].

Our work illustrates the blurred diagnostic boundaries of some neuropsychiatric disorders, such as autism and ADHD [38,39]. As an example, although the canonical Attention subscale was higher in SPARK than in ABCD, the average difference between those with ADHD versus those without was smaller in SPARK, and the association with the ADHD PGS was weaker in SPARK. Instead of replicating the canonical Attention factor in our SPARK-derived factor analysis, an alternative structure emerged that blended inattentive and atypical behaviors, which were canonically grouped into the Thought dimension. Additionally, the most clinically significant Thought problem items regarding hallucinations and self-harm were parsed into a distinct factor. This Psychoaffective Factor-6 aligns with the Hierarchical Taxonomy of Psychopathology (HiTOP) dimensions, in which thought disorder and mania dimensions are closely related and indicative of schizophrenia and bipolar spectrum disorders [40]. Thus, assessment of psychiatric symptomology in samples with elevated psychiatric symptoms and autism-specific aspects can clarify experiential and etiological overlaps among neuropsychiatric disorders.

Our study represents the most highly powered psychometric validation effort to date of psychiatric symptoms in autism. Our focus on the CBCL, one of the most well-established and widely used psychiatric questionnaires in pediatric populations, facilitates the adaptation of our findings for future research and clinical uses. Establishing a valid structure of the CBCL’s psychiatric symptoms in large autism samples is essential for the CBCL’s broad utilization in research and clinical settings. Given the lack of measurement invariance between SPARK and ABCD and the limited transferability of the canonical subscales in SPARK, our findings caution against using only the CBCL’s canonical subscales in autism.

Limitations of our study are worth consideration. Our measure of psychiatric symptoms is from parent reporting, which may not capture psychiatric symptoms that are less observable to the parent. At the same time, parent reports are an essential source of information in routine clinical assessments. Our sample included only children with verbal abilities, which may not represent psychiatric symptom profiles of autistic children with low verbal abilities. While our SPARK-derived structure had a slightly improved fit over the canonical structure, meeting acceptable fit cutoffs for SRMR and RMSEA, CFI and TLI did not meet acceptable fit cutoffs. This is likely reflective of broad phenotypic heterogeneity in autism and high CBCL item variability in SPARK. Our PGS analyses were limited to participants of European genetic ancestry. Well-powered genome-wide association studies of neuropsychiatric disorders are based on predominantly European genetic ancestry samples, and PGS estimated from these studies show reduced generalizability to other populations. This limitation influences most common variant association studies of neuropsychiatric disorders and is not specific to this study.

### Conclusions

Refining the measurement of psychiatric symptoms can help alleviate the high rates of psychiatric distress in autism. Grounding our assessments of psychiatric symptomology in large-scale efforts like this study may lead to more accurate and precise clinical indicators, including better estimates of prognosis and treatment response. This can advance monitoring of mental health and inform personalized therapies for the autism community.

## Data Availability

Approved researchers can obtain the SPARK population dataset described in this study by applying at SFARI Base (https://base.sfari.org). Data used in the preparation of this article were obtained from the Adolescent Brain Cognitive Development (ABCD) Study (https://abcdstudy.org/) held in the NIH Brain Development Cohorts Data Sharing Platform (https://www.nbdc-datahub.org/). This is a multisite, longitudinal study designed to recruit more than 10,000 children aged 9-10 and follow them over 10 years into early adulthood. The ABCD Study is supported by the National Institutes of Health and additional federal partners under award numbers:U01DA041048, U01DA050989, U01DA051016, U01DA041022, U01DA051018, U01DA051037, U01DA050987, U01DA041174, U01DA041106, U01DA041117, U01DA041028, U01DA041134, U01DA050988, U01DA051039, U01DA041156, U01DA041025, U01DA041120, U01DA051038, U01DA041148, U01DA041093, U01DA041089, U24DA041123, U24DA041147. A full list of supporters is (https://abcdstudy.org/federal-partners.html). ABCD Consortium investigators designed and implemented the study and/or provided data but did not necessarily participate in the analysis or writing of this report. This manuscript reflects the views of the authors and may not reflect the opinions or views of the NIH or ABCD Consortium investigators. The ABCD data repository grows and changes over time. The ABCD data used in this report came from Data Release Version 5.1. DOIs can be found at (https://dx.doi.org/10.15154/z563-zd24).

https://abcdstudy.org/

