## Supplemental Text and Figures for "Phenotypic structure and genetic correlates of adolescent psychiatric symptomatology in versus out of autism"

#### Supplemental Methods

##### 1. SPARK Participants and Phenotypes.

###### 1.1. Child Behavior Checklist in SPARK.

We used SPARK Phenotype Version 19, March 2026 data release (1). Our quality control of the SPARK Child Behavior Checklist (CBCL) (2) Age Version 6 to 17 Years was as follows: confirmation that individuals had an autism diagnosis based on the registration file and were between the ages of 6-17 years when the CBCL was administered, removing individuals with the CBCL invalidity flag, removing individuals that had a missing value for any of the canonical eight-factor CBCL subscales, and removing individuals that had more than 8 out of 119 missing CBCL items (per CBCL publisher recommendations). After this filtering, 22,837 individuals remained with 0.3% of CBCL items missing, which were imputed to 0.

We then merged the SPARK CBCL with the quality-controlled ABCD CBCL dataset (N=11,041) and checked for extreme outliers based on the CBCL Total score, which has a maximum possible value of 238. We tested for outliers using the check_outliers() function from the performance R package (3) with a distributional z-statistic of 3.09 (i.e., the most extreme 0.1%). From the 33,878 individuals across SPARK and ABCD, 145 were outliers, with these outlier CBCL Total scores ranging from 149 to 238. We then filtered SPARK to match the available age range of ABCD, which was 8-15 years (remaining SPARK N=14,004). Lastly, we filtered the SPARK CBCL dataset to only include individuals with verbal abilities (N=11,111). See Table S1 for additional demographic information.

###### 1.2. Verbal Ability in SPARK.

We restricted SPARK to participants with verbal abilities due to known CBCL biases in nonverbal and minimally-verbal individuals (4,5). We defined verbal abilities in SPARK as meeting the following two criteria: positive endorsement of the first item in the Social Communication Questionnaire (SCQ) (6) that asks use of short phrases or sentences and sentence-level speech at the most recent parent-reporting from the Core Descriptives file. In order to analyze how endorsement rates of CBCL items varied based on verbal ability, we also defined a subset of individuals as non/minimally verbal. Non/minimally verbal was defined as meeting the following two criteria: the SCQ was administered when the participant was at least four years old, and the first item was negatively endorsed (i.e., not able to speak words or phrases by 4 years old), and the most recent parent-reporting of language ability was either not speaking or speaking single words only.

Out of the 14,004 SPARK participants with their CBCL age range between 8-15 years, N=11,111 were defined as having verbal abilities (79.3%), N=1,198 (8.6%) as not having verbal abilities, and N=1,695 (12.1%) with an undefined verbal status due to unavailable SCQ and/or parent-reporting of language ability. The non/minimally verbal group had low endorsement rates of CBCL items requiring verbal abilities (Table S4). For example, the CBCL item “bragging, boasting” had an endorsement rate of 0.05% in the non/minimally verbal group compared to 41% in SPARK with verbal abilities and 21% in ABCD.

##### 1.3. Cognitive Impairment in SPARK.

Cognitive impairment status was determined from the parent’s most recent report on whether the participant has been diagnosed with cognitive impairment or intellectual disability. This variable was from the Core Descriptives file, and the rate in our sample was 29.2%. Additional indicators of cognitive impairment status are provided by SPARK in the Approximated Cognitive Impairment file, including the derived hierarchical cognitive impairment status (60.0%; available N=10,935) and the machine learning predicted cognitive impairment status (52.0%, available N=6,071).

##### 1.4. Co-occurring Neuropsychiatric Diagnoses in SPARK.

We used the Background Medical History Questionnaire file from SPARK to determine co-occurring diagnoses. This questionnaire is typically completed during or shortly after registering for SPARK, meaning that it is usually completed earlier than the CBCL. We therefore restricted the medical history questionnaire to participants who were at least six years old when the medical history questionnaire was administered (N=9,517). We defined anxiety disorders to include generalized anxiety disorder, social anxiety disorder, and separation anxiety disorder. Table S1 reports the prevalence of each diagnosis in SPARK and ABCD.

##### 1.5. Full-Scale IQ in SPARK.

We used the binned full-scale IQ variable from the Core Descriptives file where available (N=5,534). This variable asked the parent to select the range of the overall IQ from the most recent cognitive testing (if the participant had received cognitive testing). We redefined the IQ ranges to be equal in number on each side of the ± one SD from the mean (90-109). Therefore, the seven FSIQ ranges were: ≤69, 70 - 79, 80 - 89, 90 - 109, 110 - 119, 120 - 129, and ≥130.

#### 2. ABCD Participants and Phenotypes.

##### 2.1. Child Behavior Checklist in ABCD.

We used ABCD Version 5.1 (7), which had data collected from the baseline intake to the four-year follow-up data collection. We filtered the CBCL dataset to have no missing items (N=11,621). We then removed individuals who reported a diagnosis of autism, schizophrenia, cognitive impairment, and/or epilepsy at the baseline intake screener (3% of individuals removed). We then kept one CBCL per individual by randomly sampling across the distributions of ages (collection events). This resulted in N=11,041 individuals with their CBCL collection event sampling as follows: 25% at baseline, 16% at the one-year follow-up, 17% at the two-year follow-up, 18% at the three-year follow-up, and 24% at the four-year follow-up. The ages ranged from 8 to 15 years. Merging with the SPARK CBCL dataset and testing for extreme outliers on the CBCL Total Score did not remove any ABCD individuals (the maximum CBCL Total Score in ABCD was 131).

##### 2.2. Psychiatric Diagnoses in ABCD.

To make the psychiatric diagnoses dataset as comparable as possible with the SPARK background medical history questionnaire, we used the Kiddie Schedule for Affective Disorders and Schizophrenia (KSADS) (8) administered at baseline intake. Individuals were considered a diagnosed case if they had a past, partial remission, or current positive value for that diagnosis. The composite anxiety disorder diagnosis variable included generalized anxiety disorder, social anxiety disorder, panic anxiety disorder, agoraphobia anxiety disorder, phobia anxiety disorder, and separation anxiety disorder. The composite depressive disorder diagnosis variable included major depressive disorder, persistent depressive disorder, and unspecified depressive disorder. Table S1 reports the prevalence of each diagnosis in SPARK and ABCD. Our derived prevalences in ABCD are essentially equivalent to the prevalences reported in (9).

#### 3. Genetic Data Quality Control and Processing.

##### 3.1. Genotype Quality Control and Imputation in SPARK.

We used the common variant genotypes from SPARK integrated releases iWES1, iWES2, and iWES3. iWES1 was genotyped with the Global Screening Array, and iWES2 and iWES3 were genotyped with Twist genotyping-by-sequencing. Each iWES dataset underwent quality control and genetic ancestry inference before release by SPARK. Before imputation, we performed additional internal quality control of each iWES in parallel using PLINK (10) to follow similar steps implemented by the RICOPILI quality control pipeline (11). First, we filtered to the European genetic ancestry group (as defined by SPARK) and filtered the variants to non-ambiguous, autosomal, and single-nucleotide variants only. Second, we filtered for sample missingness <0.05, variant missingness <0.02, and minor allele frequencies $>1\times10^{-8}$ (non-founders only). Third, we filtered for sample missingness <0.02, variant missingness <0.01, and minor allele frequencies >0.001 (non-founders only), and Hardy-Weinberg equilibrium $<1\times10^{-10}$. Fourth, we filtered out trios and variants with high Mendelian error rates (max per-trio error rate 0.005 and max per-variant error rate 0.00075), and remaining low-rate Mendelian errors were set to missing. Lastly, we filtered for sample missingness <0.02, variant missingness <0.01, and minor allele frequencies >0.001 (non-founders only). After our additional internal quality control of each iWES genotype dataset, we imputed to the Haplotype Reference Consortium (HRC) reference panel with RICOPILI (11). We then restricted the variants to include only the 1.4 million HapMap3+ variants (12) and then merged the datasets across batches. Because RICOPILI reports unknown for low-confidence imputed calls, we then filtered to variant missingness <0.05 and sample missingness <0.03, leaving 1.01 million variants with a 99% genotyping rate. To make the genotypes compatible with downstream LDpred2 (12) tools that do not allow any missingness, we used the snp_fastImputeSimple function from LDpred2 with the bigsnpr R package (13) to impute these remaining calls.

##### 3.2. Genotype Quality Control and Imputation in ABCD.

ABCD was genotyped on the Smokescreen Array and released to researchers after thorough quality control and imputation to the TopMed imputation panel (14,15). We lifted over the imputed genotype files from hg38 to hg19. Similar to our SPARK genotype processing, we filtered the imputed genotypes to the 1.4 million HapMap3+ variants, leaving 1.36 million variants in ABCD. In order to filter the ABCD sample to European genetic ancestry, we ran principal component analysis (PCA) using the bigstatsr and bigsnpr R packages (13,16) with the curated unrelated participant IDs as the PCA founders, removing high LD regions and clumped SNPs. The PCs were then projected onto all ABCD participants. We then ran the n_cluster_elbow() function from the parameters R package (17) using the 20 PCs, which determined that the optimal number of clusters was 2. We then filtered the final set of ABCD participants to those whose self-reported ancestry was White and who were in the European genetic ancestry PC cluster. Lastly, we filtered the final set of ABCD variants to those that were available across all SPARK batches and ABCD (930,203 variants).

##### 3.3. Merging Genetic Datasets, Principal Components, and Relatedness Filtering.

We merged the imputed ABCD and SPARK genotypes (as opposed to meta-analysis) to have consistency across datasets and test for interactions between genotypes and autism diagnosis. After imputation and post-imputation quality control, we merged the genotypes and removed any variants with a minor allele frequency >0.005. Our final merged dataset consisted of 930,203 HapMap3+ variants in N=103,327 individuals of European genetic ancestry (SPARK N=97,275 and ABCD N=6,052). We ran PCA using the bigstatsr and bigsnpr R packages (13,16) with the SPARK parents and curated unrelated ABCD participants as founders, removing high LD regions and clumped SNPs. The PCs were then projected onto all SPARK and ABCD participants. We created a genetic relationship matrix using GCTA (18) and pruned related participants using GCTA's recommended relatedness cutoff of 0.05. This final sample of unrelated individuals (N=9,530; SPARK N=4,963; ABCD N=4,567) was used for the common genetic variation analyses, including SNP-based heritability and polygenic scores associations.

##### 3.4. SNP-Based Heritability.

SNP-based heritabilities ($h^2$) of the CBCL subscales were calculated using GCTA-REML (19) with age, sex, and the first five genetic principal components as covariates. The heritabilities were calculated separately in the unrelated individuals in SPARK (N=4,963) and ABCD (N=4,567). See Table S13 for all GCTA-REML $h^2$ statistics.

##### 3.5. Polygenic Scores.

We calculated polygenic scores (PGS) with LDpred2-auto (12) using the merged ABCD and SPARK, quality-controlled genotypes. We used the external LD matrix calculated from UK Biobank samples recommended in (20) and available here (21). We calculated PGS for cognitive performance (22) and four psychiatric case-control PGS: autism (23), ADHD (24), depression (25), and schizophrenia (26). We adjusted the raw PGS for the first five principal components by linear regression residualization and then standardized in our sample. We performed two validation tests of the PGS in the merged PGS. We validated that the mean PGS differences between SPARK and ABCD were as expected. We also confirmed that correlations between PGS were as expected (i.e., positive genetic correlations within psychiatric PGS) and that correlations between PGS were similar when calculated separately in SPARK versus ABCD (Table S11). PGS associations with the CBCL subscales were calculated with negative binomial regression with age and sex as covariates.

##### 3.6. De Novo Genetic Burden.

We analyzed de novo burden associations to the CBCL subscales in the subset of SPARK with complete trios (N=2,474). We defined de novo burden as carriers of a de novo protein-truncating variant or de novo coding copy-number variant in an autism-associated gene (253 genes), or carriers of a de novo genomic disorder deletion or duplication associated with autism or intellectual disability (e.g., 22q11.2 deletion). The de novo calls and autism-associated gene list are from the latest unpublished exome sequencing aggregation effort by the Autism Sequencing Consortium (27). The overall de novo burden carrier rate was 3.9%, in which 2.3% carried a protein-truncating variant, 0.6% carried an exonic copy-number variant, and 1.1% carried a genomic disorder variant (see Table S12). De novo burden carrier associations with the CBCL subscales were calculated with negative binomial regression with age and sex as covariates.

#### 4. Statistical analyses.

##### 4.1. Factor Analyses.

As all CBCL items are on a 3-point Likert scale, we used factor analysis methods with ordinal specifications. Confirmatory factor analyses (CFAs) were conducted using the cfa() function from the lavaan (28) R package using the weighted least squares mean-and-variance-adjusted (WLSMV) estimator: cfa(model, data, estimator="WLSMV", ordered=TRUE, std.lv=TRUE, parameterization="delta").

We assessed CFA measurement invariance across ABCD versus SPARK groups using multigroup CFA. We tested increasingly restrictive levels of invariance, including configural invariance (same factor structure across groups), metric invariance (equal factor loadings across groups), and scalar invariance (equal factor loadings and item intercepts across groups).

CBCL item means and variances were higher in SPARK (average mean of 0.52; average variance of 0.40) than in ABCD (average mean of 0.14; average variance of 0.14; Table S4). We removed the following five items that had near-zero variance in both cohorts: drinks alcohol, uses tobacco, uses drugs, sets, fires, and sexual problems. With the remaining 114 items, we used three methods to determine the optimal number of factors to extract in SPARK: 1) variance explained from principal component analysis of the correlation matrix, 2) scree $R^2$ (29), and 3) sequential exploratory factor analysis (EFA) from one to 15 factors (Table S5). We determined a seven-factor solution to be optimal based on each component explaining greater than 2% of the variance, as well as 47% cumulative variance explained and a scree $R^2$ of 0.77 (recommended cutoff ≥0.75). EFAs were conducted using the psych (30) R package with Promax rotation, weighted least squares factoring method, and polychoric correlations. We compared EFA structures using the Tucker congruency coefficient method (31) (φ), in which φ greater than 0.95 indicates nearly identical factors.

The final SPARK-derived EFA was converted to a CFA structure and item-summed subscales using a loading threshold of ≥0.3 (Figure S2). Due to the possibility of an item having more than one meaning and/or a different meaning in the context of autism (e.g., running away from home potentially corresponding to either rule-breaking or elopement in autism due to sensory overwhelm (32)), we allowed items to cross-load to more than one factor. The final SPARK-derived factor structure leveraged 103 items, 20 of which cross-loaded to two factors and one item that cross-loaded to three factors (Table S7).

**4.2. Phenotypic and Genetic Associations with the CBCL Subscales.**
All phenotypic and genetic associations to the CBCL subscales included sex and age at CBCL assessment as covariates. Standardized coefficient estimates with 95% confidence intervals were extracted from models using the parameters R package (17). We used logistic regression for case/control diagnosis outcomes with the glm() R function, ordinal logistic regression for full-scale IQ outcomes with the polr() function from the MASS R package (33), and negative binomial regression for CBCL subscale outcomes using the glm.nb() function from the MASS R package (33). For t-tests and correlations, the CBCL subscales were adjusted for age and sex with negative binomial regression residualization and then standardized. P-values were adjusted for multiple comparisons using the false discovery rate (FDR) method unless stated otherwise with $p_{nominal}$. P-values were adjusted separately per cohort and CBCL subscale group (i.e., canonical and SPARK-derived) because of redundancy between subscale groups and to maintain the same adjustment level between cohorts.

### Supplemental Figures

**Figure S1: Stability of the Seven-Factor CBCL structure in SPARK.**
We split the SPARK sample randomly into two sets and assessed the congruency between EFA structures. The congruency (Tucker congruency coefficient, φ) between factor pairs are listed below factor names.

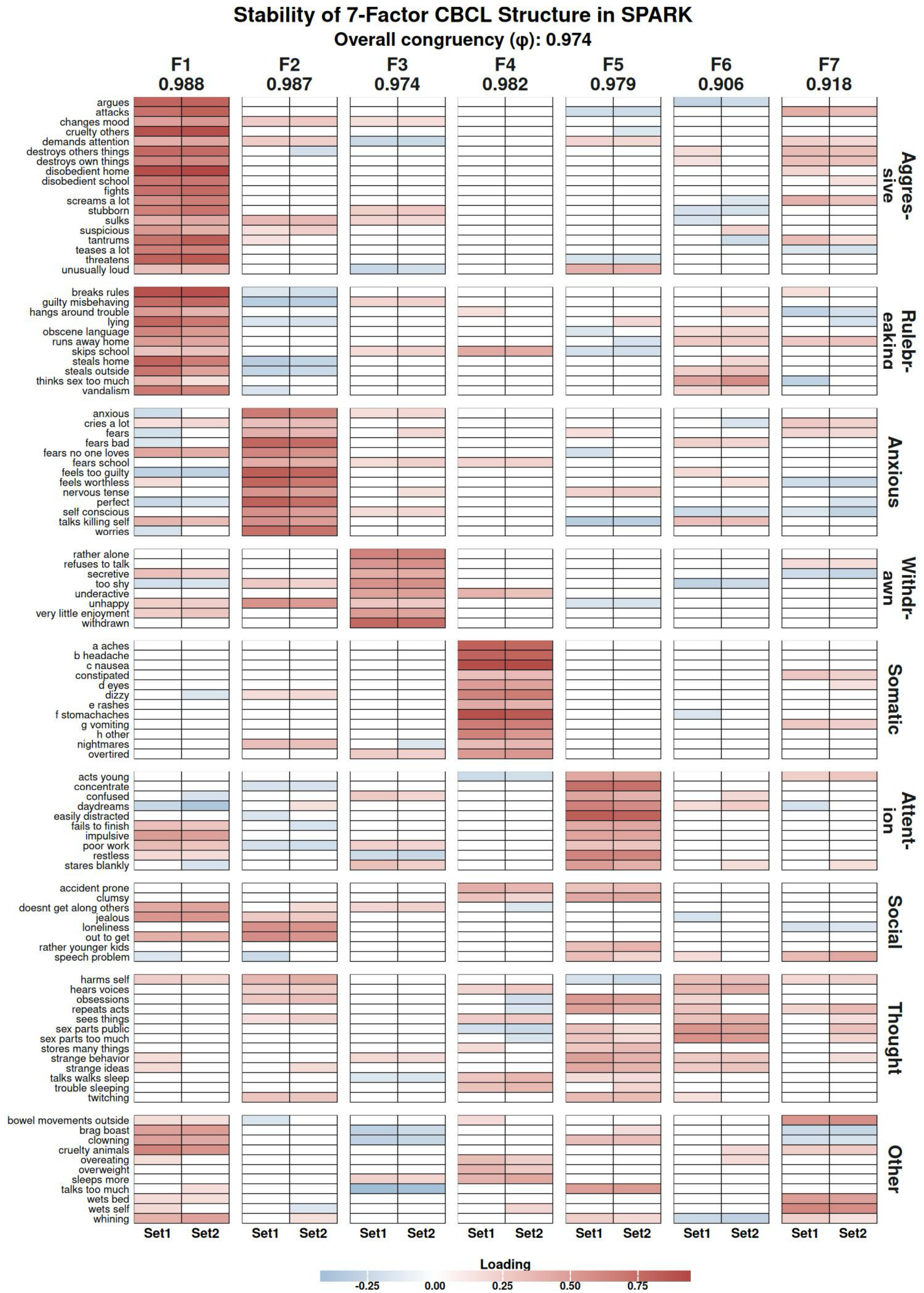

**Figure S2: Distributions of the SPARK-derived-7F CBCL subscales.**
Distributions of the SPARK-derived-7F CBCL subscales in SPARK (N=11,111).

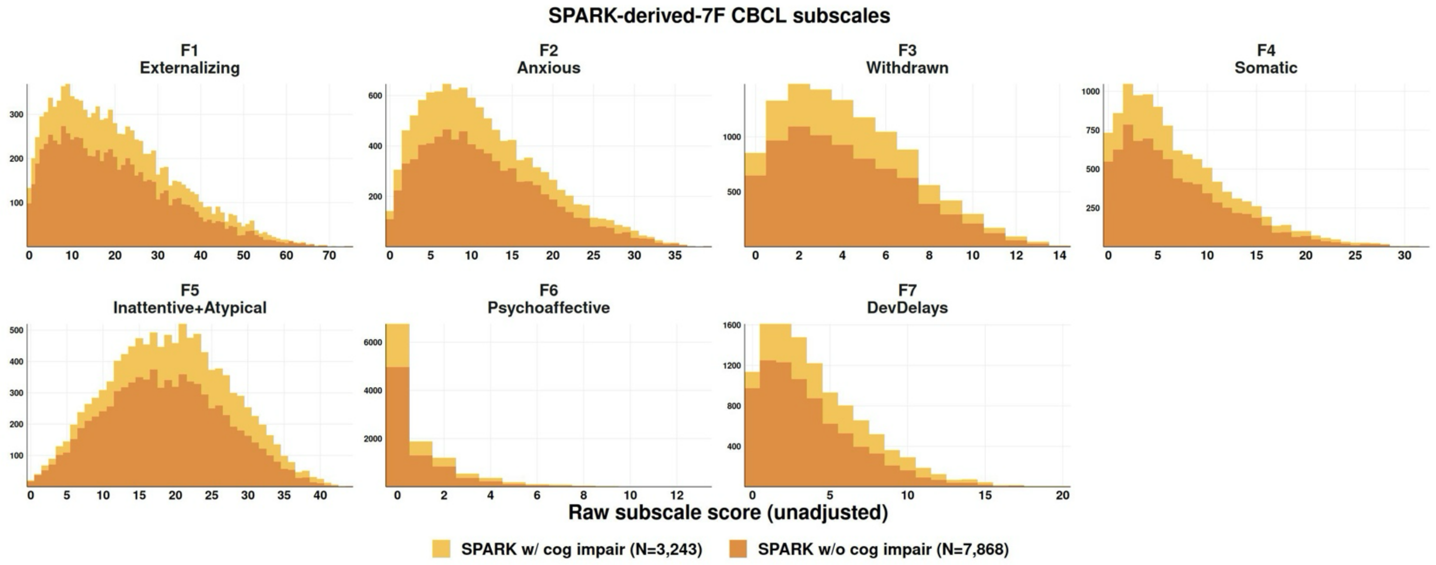

### List of Supplemental Tables

*See Excel file for Supplemental Tables.*

**Table S1: Sample demographics.**
The statistics are mean ± SD (minimum, maximum) for the continuous variables and N with % of total N for the categorical variables. Sample sizes are reported with variables that were not available in 100% of the sample. Race/ethnicity is the parent/guardian reporting at registration and sex is sex at birth.

**Table S2: Mean differences in the Canonical 8-Factor CBCL subscales by cohort, cognitive impairment, verbal ability, and genotyped versus not-genotyped groups.**
T-tests and Wilcox tests with phenotypes adjusted for age and sex then standardized.

**Table S3: Associations between four psychiatric diagnoses and the Canonical 8-Factor CBCL subscales in SPARK versus ABCD.**
Logistic regression with age and sex as covariates and t-tests with the CBCL subscales adjusted for age and sex then standardized.

**Table S4: Rates of endorsement, means, and variances of each CBCL item.**
Rates of endorsement (Likert 1 or 2) are reported for SPARK (verbal) and ABCD, as well as the non/minimally verbal SPARK sample (N=1,198) that was not included in the main analyses.

**Table S5: Determining the optimal number of factors to extract from the CBCL in SPARK.**
We tested 1-15 factors using three methods: 1) variance explained from principal component analysis of polychoric correlation matrix of the CBCL itemset, 2) scree $R^2$, and 3) sequential EFA.

**Table S6: Factor analyses metrics for Canonical 8-Factor, SPARK-Derived 7-Factor, and ABCD-Derived 7-Factor CBCL structures.**
Confirmatory factor analysis (CFA), CFA measurement invariance, exploratory factor analysis (EFA), and EFA g-factor model metrics for SPARK and ABCD subgroups.

**Table S7: SPARK-Derived factor metrics and loadings.**
EFA variance explained, factor congruency (φ) with ABCD-Derived 7-Factor, EFA factor loadings, EFA item complexity and uniqueness, and the list of items retained per factor in lavaan CFA syntax. EFA factor loadings ≥ 0.3 were retained for the conversion of EFA to CFA and in calculations of the SPARK-Derived CBCL subscales.

**Table S8: Correlations between CBCL subscales in SPARK versus ABCD.**
Pearson correlation coefficients with 95% CI.

**Table S9: Associations between full-scale IQ and CBCL subscales in SPARK.**
Ordinal logistic regression with age and sex as covariates, Kruskal tests, and mean CBCL subscale score with 95% CI per full-scale IQ group. The CBCL subscales were adjusted for age and sex and then standardized before Kruskal tests and calculating group means.

**Table S10: Associations between eight co-occurring neuropsychiatric diagnoses and CBCL subscales in SPARK.**
Logistic regression with age and sex as covariates. T-tests and Wilcox tests with phenotypes adjusted for age and sex then standardized.

**Table S11: Polygenic score cohort comparisons.**
Polygenic score (PGS) mean differences between SPARK versus ABCD. Correlations between PGS pairs were calculated separately per cohort.

**Table S12: Rates and type of de novo burden in SPARK.**
Rates of overall de novo burden carrier status as well as rates for the variant type (protein-truncating variant, copy-number variant, and/or genomic disorder).

**Table S13: SNP-based heritabilities of CBCL subscales in SPARK versus ABCD.**
SNP-based heritabilities ($h^2$) with standard error (SE), and the log-likelihood ratio test statistic (LRT) from GCTA-REML.

**Table S14: Polygenic score and de novo burden associations with the CBCL subscales in SPARK versus ABCD.**
Negative binomial regressions were performed separately per cohort and in combined-cohort models with a PGS-by-cohort[SPARK] interaction term.
